# Clinical Validation of Metabolite Markers for Early Lung Cancer Detection

**DOI:** 10.1101/2024.11.25.24317901

**Authors:** Lun Zhang, Jiamin Zheng, Rashid Ahmed, Jeff Haince, Claudia Torres-Calzada, Rupasri Mandal, Andrew Maksymuik, Paramjit S. Tappia, Philippe Joubert, Christian D. Rolfo, David S. Wishart

## Abstract

Non-small cell lung cancer (NSCLC), comprising 85% of lung cancers, is a leading cause of cancer mortality. Early detection enhances survival, but current screening methods are limited. This retrospective study used targeted mass spectrometry-based metabolomics on 680 plasma samples from NSCLC patients and controls (discovery cohort) and 216 samples (validation cohort). Logistic regression models with a subset of ten metabolites achieved over 90% area under the ROC curve (AUROC) for distinguishing patients from controls, including early-stage disease. Incorporating smoking history improved model performance. In the discovery cohort, AUROCs were 93.6% (all stages), 93.7% (Stage I and II), and 93.9% (Stage I). Validation confirmed the high sensitivity and specificity of the models. This study demonstrates that metabolomic biomarkers provide a minimally invasive, sensitive, and specific tool for early NSCLC detection, potentially improving screening and patient outcomes. Future studies should validate these biomarkers in diverse populations.

**Statement of significance:** This study identifies plasma metabolite biomarkers that enable sensitive and specific early detection of NSCLC using minimally invasive blood sampling. Achieving over 90% area under the ROC curve for early-stage patients, the findings promise to improve lung cancer screening methods and enhance early interventions and patient outcomes.

## Introduction

Lung cancer ranks among the primary causes of cancer-related fatalities worldwide. It is a complex disease that arises from the uncontrolled growth of abnormal cells in lung tissue. There are two main types of lung cancer: non-small cell lung cancer (NSCLC) and small cell lung cancer (SCLC). NSCLC accounts for approximately 85% of all lung cancer cases and is further classified into three subtypes: adenocarcinoma, squamous cell carcinoma, and large cell carcinoma (1).

According to the American Cancer Society and the Canadian Centre for Applied Research in Cancer Control, lung cancer is among the two most commonly diagnosed cancers in these countries, and the leading cancer-related death in both countries (2,3). The diagnosis and treatment of lung cancer are challenging due to its diverse subtypes, varying stages, spatial-temporal heterogeneity, and complex biological mechanisms.

Early detection of lung cancer is crucial for improving patient outcomes, as early diagnosis is associated with higher survival rates. Patients with stage IA lung cancer have a high 5-year survival rate exceeding 75%, but^5^ as cancer progresses, the long-term survival rate dramatically decreases (4). Over the years, various methods have been developed for detecting lung cancer, ranging from traditional imaging techniques to cutting-edge molecular tests. The most common methods to detect lung cancer include imaging techniques such as chest X-rays, computed tomography (CT) scans, and positron emission tomography (PET) scans. These methods can help detect lung nodules or tumors and determine their size and location. However, imaging tests may not always provide a definitive diagnosis, and further testing such as lung biopsies may be required. Molecular tests such as liquid biopsies, which detect circulating tumor DNA (ctDNA) or other biomarkers in blood samples, and genetic testing of lung cancer tissue, can also aid in the diagnosis and management of lung cancer (5).

The application of low-dose computed tomography (LDCT) in the context of lung cancer screening programs provide the possibility of detecting lung cancer at earlier, more operable stages in high-risk populations. However, LDCT by itself is a limited approach, with lower sensitivity and specificity compared with regular chest CT scan (6,7).

In recent years, metabolomics has emerged as a promising tool in the field of cancer research, including lung cancer. Metabolomics is the comprehensive study of metabolites, which are small molecules produced by cellular processes. Metabolomics not only provides a snapshot of the metabolic state of a cell or organism (8), but it can also be used to identify biomarkers for early prediction and diagnosis of lung cancer, reveal the pathogenesis of the disease, and provide insights into potential therapeutic targets (9–12). By analyzing the metabolic profiles of biological samples, such as blood, plasma, or urine, metabolomics can detect changes in metabolite levels that are associated with lung cancer (13). This approach has the potential to improve the accuracy and sensitivity of lung cancer detection, leading to the development of more effective diagnostic strategies. In this context, metabolomics holds great promise for improving the detection, diagnosis, and treatment of lung cancer. However, further research is needed to validate metabolomic biomarkers and identify their clinical utility in the management of this devastating disease.

The purpose of this study is to build upon our previous research (14) and to identify a more robust panel of plasma metabolites for early-stage lung cancer diagnosis by using a much larger and more complex patient cohort. While earlier studies (13, 14) have demonstrated the promise of metabolomics in distinguishing lung cancer patients from healthy individuals, further validation and identification of a more reliable set of biomarkers in a broader and more diverse cohort is essential. To achieve this, we employed the same quantitative MS-based metabolomics assay used in (14) to analyze a significantly larger set of plasma samples from both lung cancer patients and non-cancer controls (many with other lung diseases). The resulting high-performing biomarker panel was then validated on a slightly smaller cohort with a similarly complex patient structure and found to exhibit essentially the same high diagnostic performance. This work has the potential to improve the clinical utility of metabolomics in the early detection and diagnosis of lung cancer, ultimately leading to better patient outcomes.

## Materials and Methods

### Regulatory and Institutional Review Board Approvals

Ethics approval was obtained from the University of Manitoba Health Research Ethics Board (Ethics File #: H2012:334) prior to the implementation of the study. Research ethics approval was also obtained from the University of Alberta (Study ID: Pro00093715) to conduct the metabolomic studies in Edmonton.

### Study Population and Sample Collection

All plasma samples were obtained from the IUCPQ (Institut Universitaire de Cardiologie et de Pneumologie de Quebec) Biobank-Respiratory Healthy Research Network. Frozen (-80 °C) plasma aliquots ranging from 200 to 400 µL were shipped to The Metabolomics Innovation Centre (TMIC) at the University of Alberta, Canada for quantitative metabolomic analysis. The 680 archived plasma samples that were used as the discovery cohort in this study were collected from 466 patients with biopsy-proven and biopsy-graded lung cancer and 214 controls. The 216 plasma samples that were used as the validation set were collected from 156 patients with biopsy-proven and biopsy-graded NSCLC and 60 controls (15). The cancer samples had comprehensive data including demographics, body mass index (BMI), smoking status, overall survival, morbidities, pathology, etc. The control samples had data on demographics, BMI, and medical condition history. Patients and controls with a history of any liver or kidney disease, and any previous treatment with anti-neoplastic drugs were excluded from this cohort. In addition to healthy individuals, the control groups also included patients with various pulmonary conditions such as asthma, chronic obstructive pulmonary disease (COPD), bronchiectasis, hamartoma, and COVID. This was done to investigate whether the discovered metabolite markers were specific to lung cancer alone rather than lung disease. More information about the control groups is summarized in Table S1.

### Chemicals, Reagents, and Materials Used for the Quantitative Metabolomic Assays

Pure reference standard compounds used for the quantitative metabolomics analysis, Optima™ LC-MS grade ammonium acetate, phenylisothiocyanate (PITC), 3-nitrophenylhydrazine (3-NPH), 1-ethyl-3-(3-dimethylaminopropyl) carbodiimide (EDC), HPLC grade pyridine, HPLC grade methanol, HPLC grade ethanol, and HPLC grade acetonitrile (ACN) were purchased from Sigma-Aldrich (Oakville, ON, CA). Optima™ LC-MS grade formic acid and HPLC grade water were purchased from Fisher Scientific (Ottawa, ON, CA). ^2^H-, ^13^C- and ^15^N-labelled compounds were purchased from Cambridge Isotope Laboratories, Inc. (Tewksbury, MA, USA) and Sigma-Aldrich (Oakville, ON, CA). 3-(3-hydroxyphenyl)-3-hydroxypropionic acid (HPHPA) and ^13^C-labelled HPHPA were synthesized in-house as described previously (16). Multiscreen “solvinert” filter plates (hydrophobic, PTFE, 0.45 μm, clear, non-sterile) and Nunc^®^ 96 DeepWell™ plates were purchased from Sigma-Aldrich (Oakville, ON, CA).

### Stock Solutions, Internal Standard (ISTD) Mixture, and Calibration Curve Standards for Metabolomic Assays

All chemicals, including isotope-labelled compounds, used in this study were weighed individually on a Sartorius CPA225D semimicro electronic balance (Mississauga, ON, CA) with a precision of 0.0001 g. Stock solutions with proper concentrations for each analyte, and stock solutions of internal standards (ISTD) were prepared by dissolving the accurately weighed chemicals in proper solvents. Calibration curve standards, quality control standards, and working ISTD solutions were prepared by mixing and diluting corresponding stock solutions with appropriate solvents. All standard and ISTD solutions were aliquoted and stored at –80 °C until further use.

### Sample Preparation and Liquid Chromatography/Direct Injection Mass Spectrometry for Metabolomic Assays

The plasma samples were analyzed using the same targeted, quantitative MS-based metabolomics approach as in our previous lung cancer biomarker study (14). Mass spectrometric analysis of the diluted plasma extracts was performed on a Qtrap^®^ 4000 tandem mass spectrometry instrument (Applied Biosystems/MDS Analytical Technologies, Foster City, CA) equipped with an Agilent 1260 HPLC (Agilent Technologies, Santa Clara, CA, USA). This LC-MS assay enables the targeted identification and quantification of up to 138 different endogenous metabolites, including amino acids, acylcarnitines, biogenic amines and derivatives, organic acids, uremic toxins, glycerophospholipids, sphingolipids, and sugars. The method employs chemical derivatization (via 3-NPH for organic acids or PITC for amine-containing compounds), analyte extraction, analyte separation, and selective mass-spectrometric detection using multiple reaction monitoring (MRM) pairs for metabolite identification and quantification. Isotope-labeled ISTDs, along with other ISTDs are used for accurate metabolite quantification. Details of the method, derivatization strategy, separation protocol, MS methods, calibration, and metabolite quantification process are described in (17) .

### Data Analysis

Numeric clinical variables were analyzed using a Student’s t-test or Mann–Whitney rank sum test, depending on their normality. The normality of the numerical clinical variables was analyzed by a Shapiro-Wilk test. Categorical clinical variables were analyzed by χ^2^ tests. All these tests mentioned above were performed with a p-value threshold at 0.05 using the R statistical programming language (R 4.2.1) (18). Recommended statistical procedures for standard quantitative metabolomic analysis were followed (19). Metabolites with >80% missing values across all groups were removed from further analysis. For metabolites with less than 80% missing concentrations, values were imputed by using 1/5 of the minimum detectable concentration value for that metabolite. The raw concentrations underwent median normalization, log transformation, and then auto-scaling (mean-centered and divided by the standard deviation of each variable) before further data analysis. Non-parametric univariate analysis and partial least squares discriminant analysis (PLS-DA) were performed by using MetaboAnalyst 5.0 (20). A 1000-fold permutation test was conducted to determine the likelihood that the observed separation of the PLS-DA was not due to chance. Predictive models for lung cancer were developed using logistic regression, incorporating both metabolite and clinical variables. Logistic regression was performed by using R 4.2.1 (18). Optimal regression models were first identified using the discovery cohort. These models were then confirmed using the validation cohort. The area under the receiver-operator characteristic curves (AUROC), sensitivities/specificities at selected cut-off points, and the 95% confidence intervals were calculated for both the discovery and the validation sets for all models using the pROC R package (21). Cut-off points were determined by calculating the Youden Index (J = max {Sensitivity + Specificity − 1}).

## Results

### Clinical Cohorts

A summary of the clinical variables for both the discovery set and the validation set is listed in Table 1. The results from a comparison of the different sets can be found in Table S1. Comparisons of weight and BMI between each cancer group and the control group revealed no statistically significant differences. The χ^2^ test results showed that the cancer groups and the controls were well matched in terms of sex. However, the Mann-Whitney rank sum tests highlighted a significant age difference between the controls and each cancer group. Despite the minor fold-change in age between the controls and each cancer group (for instance, the age fold-change of Stage I and Stage II versus controls is 1.06 and 1.08, respectively), the significance impact of age was assessed by including it as a covariate in the subsequent modeling phase.

**Table 1.**
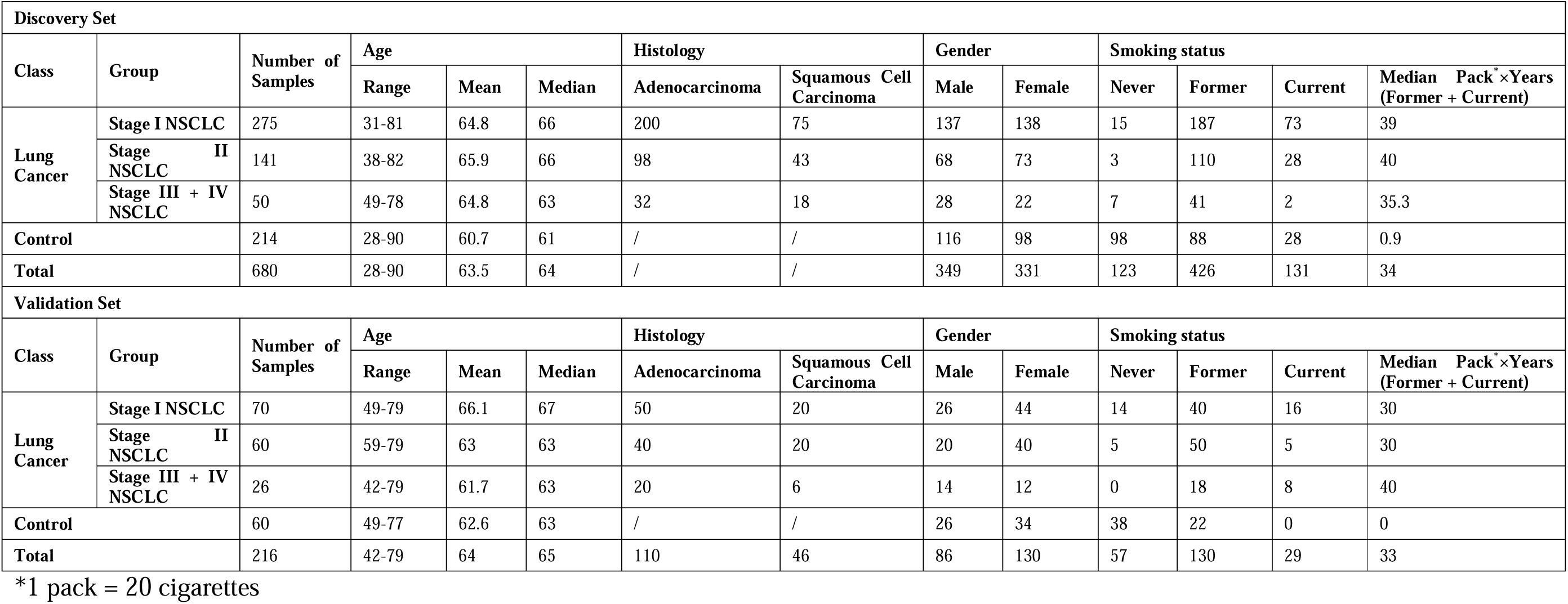
Summary of grouping of samples.

| Discovery Set |  |  |  |  |  |  |  |  |  |  |  |  |  |
| --- | --- | --- | --- | --- | --- | --- | --- | --- | --- | --- | --- | --- | --- |
| Class | Group | Number of Samples | Age |  |  | Histology |  | Gender |  | Smoking status |  |  |  |
|  |  |  | Range | Mean | Median | Adenocarcinoma | Squamous Cell Carcinoma | Male | Female | Never | Former | Current | Median Pack*×Years (Former + Current) |
| Lung Cancer | Stage I NSCLC | 275 | 31-81 | 64.8 | 66 | 200 | 75 | 137 | 138 | 15 | 187 | 73 | 39 |
|  | Stage II NSCLC | 141 | 38-82 | 65.9 | 66 | 98 | 43 | 68 | 73 | 3 | 110 | 28 | 40 |
|  | Stage III + IV NSCLC | 50 | 49-78 | 64.8 | 63 | 32 | 18 | 28 | 22 | 7 | 41 | 2 | 35.3 |
| Control |  | 214 | 28-90 | 60.7 | 61 | / | / | 116 | 98 | 98 | 88 | 28 | 0.9 |
| Total |  | 680 | 28-90 | 63.5 | 64 | / | / | 349 | 331 | 123 | 426 | 131 | 34 |
| Validation Set |  |  |  |  |  |  |  |  |  |  |  |  |  |
| Class | Group | Number of Samples | Age |  |  | Histology |  | Gender |  | Smoking status |  |  |  |
|  |  |  | Range | Mean | Median | Adenocarcinoma | Squamous Cell Carcinoma | Male | Female | Never | Former | Current | Median Pack*×Years (Former + Current) |
| Lung Cancer | Stage I NSCLC | 70 | 49-79 | 66.1 | 67 | 50 | 20 | 26 | 44 | 14 | 40 | 16 | 30 |
|  | Stage II NSCLC | 60 | 59-79 | 63 | 63 | 40 | 20 | 20 | 40 | 5 | 50 | 5 | 30 |
|  | Stage III + IV NSCLC | 26 | 42-79 | 61.7 | 63 | 20 | 6 | 14 | 12 | 0 | 18 | 8 | 40 |
| Control |  | 60 | 49-77 | 62.6 | 63 | / | / | 26 | 34 | 38 | 22 | 0 | 0 |
| Total |  | 216 | 42-79 | 64 | 65 | 110 | 46 | 86 | 130 | 57 | 130 | 29 | 33 |

\*1 pack = 20 cigarettes

Furthermore, it is important to note that there are differences in the smoking status between the cancer groups and the control group, as outlined in Table 1. The correlation between smoking history and lung cancer is a topic has been extensively researched and is widely recognized. In our dataset, a very significant association between smoking history and the incidence of lung cancer was observed. For instance, when considering all cancer patients as a single group, the *p*-value of the χ^2^ test was 2.2 × 10^-16^. Moreover, both sets exhibited notably higher medians of smoking amount (measured in packs × years) in the cancer groups compared to the control groups. As a result, the smoking status was included as a cofactor in the subsequent modeling due to its relevance and potential influence.

### Univariate and Multivariate Statistical Analysis

In our previous study (14), a significant distinction between the metabolomics profile of patients with NSCLC and healthy individuals was discovered. In this study, a similar set of analyses was conducted initially for all NSCLC patients and controls. All metabolic features in the dataset were processed as outlined in the Materials and Methods section. These analyses aimed to explore the alterations in the metabolomics profiles of the patients and to identify potential biomarkers associated with the disease.

The PLS-DA analysis revealed distinct metabolomics profiles between patients with NSCLC and control individuals (Fig 1A). The permutation test confirmed that the class discrimination is significant (Fig S1A). In the exploratory receiver-operating characteristic (ROC) analysis, based on random forest, the area under the ROC curve (AUROC) of different models, each with different number of metabolite features, ranged from 0.80 to 0.90 (Fig S2A). These results suggest that metabolic features may effectively discriminate NSCLC patients from controls. The heatmap showed that patients with NSCLC had higher plasma concentrations of acyl-carnitines and beta-hydroxybutyric acid, but lower levels of lysophosphatidylcholines (lysoPCs), citric acid, pyruvic acid and tryptophan levels, compared to the controls (Fig 1B). The Mann-Whitney rank sum tests showed that 41 out of the 138 quantitatively measured metabolites displayed significant differences between the patients with NSCLC and the controls. Both univariate analyses (using the Mann-Whitney rank sum tests) and multivariate analyses (using the PLS-DA and the random forest ROC analysis) consistently identified the same metabolites with relevant alterations. The metabolites that increased most notably were β-hydroxybutyric acid, citrulline, carnitines (carnitine and acetyl-carnitine), and succinic acid. Conversely, the metabolites that decreased most significantly included citric acid, tryptophan, various lysoPCs (lysoPC a C18:2, lysoPC a C18:0, and lysoPC a C16:0), and PC ae C40:6 (Fig S1B, S2B, S3A and Table S2).

**Fig 1.**
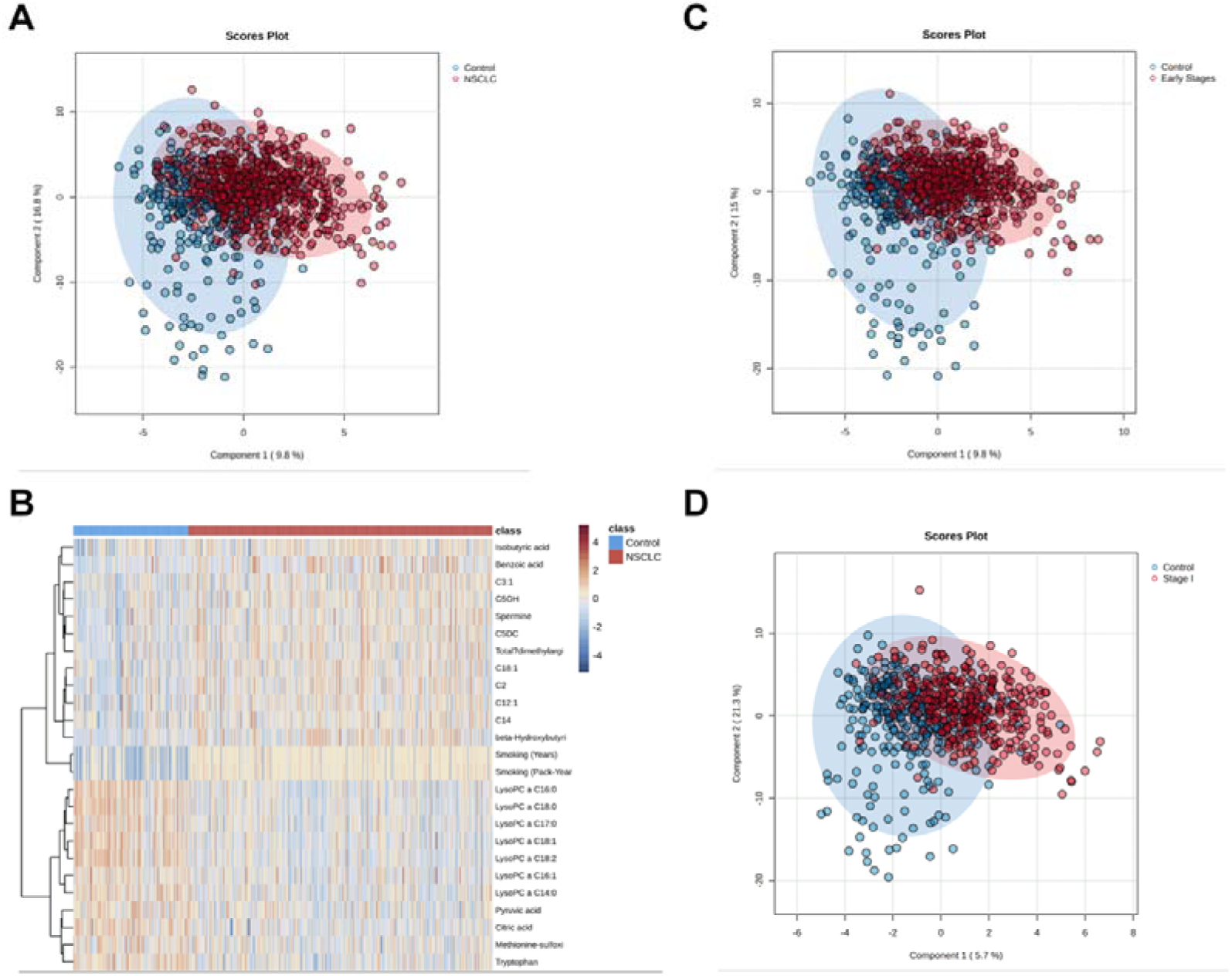
Metabolomics profiles of lung cancer patients and controls are significantly different. (A) PLS-DA 2D-scores plot of all-stages lung cancer patients vs controls. (B) A hierarchical clustering heat map of the metabolites measured in the plasma of lung cancer patients and healthy controls. Only the top 25 metabolites were shown. (C-D) PLS-DA 2D-scores plot of Early stages (stages I + II) lung cancer patients (C), and Stage I lung cancer patients (D) vs. controls.

The earlier a set of biomarkers can diagnose a condition, the more valuable it becomes. Therefore, we sequentially investigated the metabolomics profiles of patients at the early stages (Stage I + II; n=416) and at the earliest stage (Stage I; n=275), following the same analysis workflow as described above. The PLS-DA results demonstrated a separation between the controls and the patients at different disease stages (Fig 1C and 1D). The permutation test confirmed that the observed PLS-DA results were not due to chance (Fig S1B and S1C). The AUROC of different random forest-based models, with varying numbers of metabolite features, ranged from 0.81 to 0.90 (Fig S2C) for the early-stage patients, and from 0.82 to 0.91 (Fig S2E), for the stage I patients.

For both the early-stage patients (Stage I + II) and the Stage I patients alone, a similar combination of biomarkers was suggested by the PLS-DA (Fig S1D and S1F), the random forest-based ROC model (Fig S2D and S2F), and the Mann-Whitney rank sum tests (Fig S3B and S3C, Table S3 and S4). Considering that approximately 90% of the enrolled NSCLC patients were at Stages I and II, these results suggest that patients with NSCLC at Stage I and Stage II shared a similar metabolomics profile. In fact, the PLS-DA was unable to distinguish patients with NSCLC at different stages (Fig S4).

### Logistic regression modeling

Logistic regression (LR) was carried out to develop a diagnostic model of NSCLC with potential clinical utility. Initially, attention was focused solely on the metabolite features. It was suggested by the previous random forest-based ROC model exploration that high discriminative power (AUC > 85%) could be achieved by introducing no more than 15 metabolomic biomarkers (Fig S2). Multiple LR models for all stages of NSCLC, early-stages, and stage I were built and optimized using the discovery set. A combination of 10 metabolites out of an initial set of 138 was consistently identified throughout the modelling process that yielded consistent results in terms of diagnostic performance across different cancer stages. These metabolites included citric acid, tryptophan, lysoPC a C18:2, lysoPC a C20:3, carnitine, glutamine, citrulline, succinic acid, PC aa C38:0, and PC ae C40:6. Nearly identical performance in both the discovery and the validation sets was achieved by the diagnostic model. When all patients with NSCLC were included, the AUROC values for the discovery set and the validation set were 91% and 89%, respectively. When applied to early-stage patients and Stage I patients, the discovery-AUROC/validation-AUROC values were 91%/88% and 91%/85%, respectively (Fig 2A - 2C).

**Fig 2.**
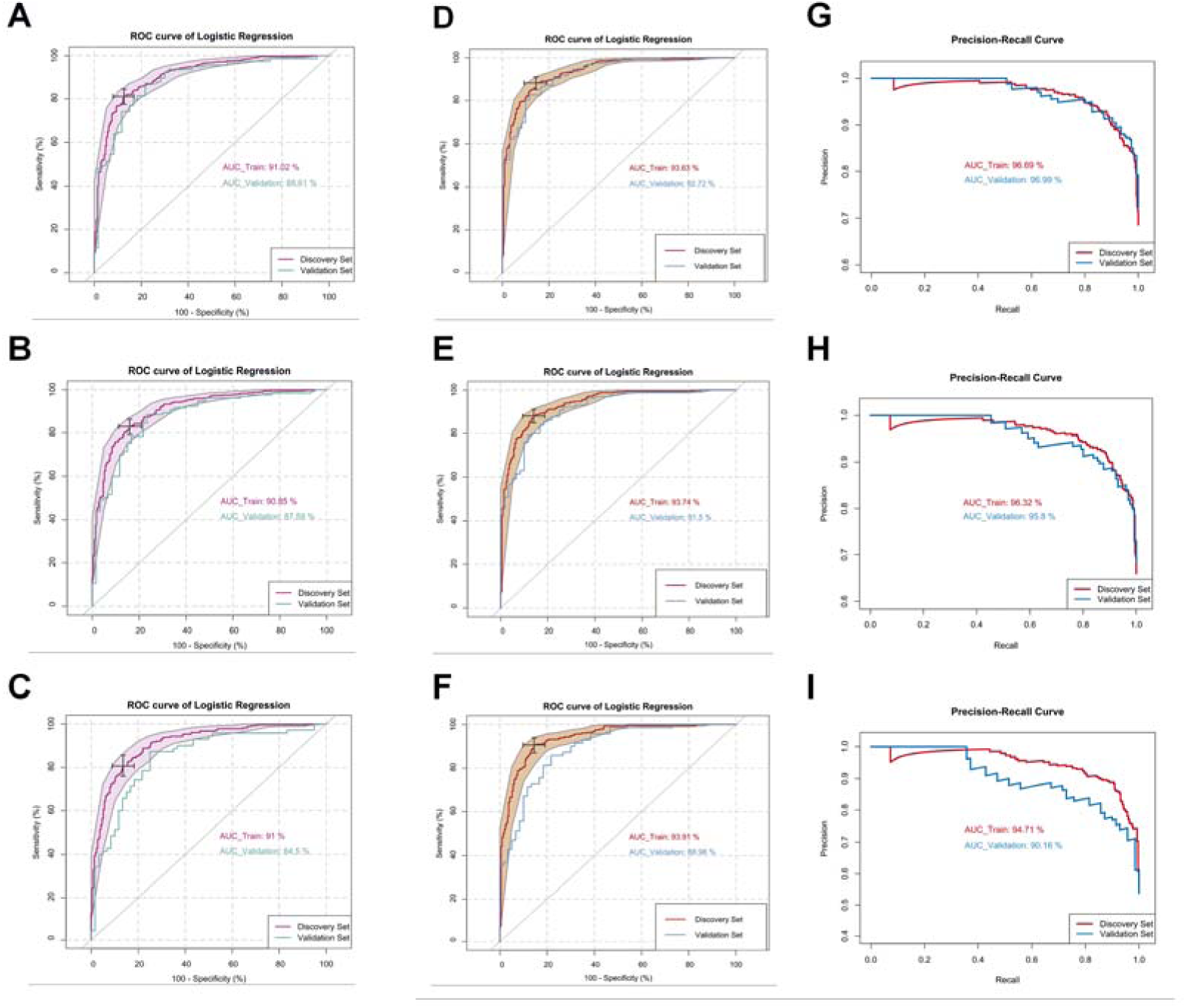
Logistic regression modeling can effectively discriminate lung cancer patients from controls. (A-C) ROC curves generated by the logistic regression models using metabolite features only for lung cancer patients at all stages (A), early stages (stage I + stage II) (B), and stage I (C). ROC curves and their 95% CI on the discovery set are shown in magenta. ROC curves obtained from the validation set are colored in cyan. (D-F) ROC curves generated by the logistic regression models using both metabolite features and smoking factor for lung cancer patients at all stages (A), early stages (stage I + stage II) (C), and stage I (E). ROC curves and their 95% CI on the discovery set are shown in red. ROC curves obtained from the validation set are colored in blue. (G-I) Precision-recall curves of the logistic regression models for lung cancer patients at all stages (B), early stages (stage I + stage II) (D), and stage I (F). Curves of the discovery set and the validation set are colored in red and blue, respectively.

As previously noted, an imbalance was observed in age and smoking amount between the NSCLC and the control groups. The significance of these potential cofactors was determined by examining their influence during the modeling process. When age alone was used to build the logistic regression model, an AUROC value of 57% was achieved, with a corresponding *p*-value of 0.61. When age was added to the previously described LR model, a *p*-value of 0.33 was obtained and the AUROC value was only increased by 0.07%. These findings suggest that age does not play a significant role in discriminating between NSCLC patients and controls. In contrast, when only the smoking amount (packs×years) was considered, an AUROC value of 82% was achieved by the logistic regression model. A p-value associated with the smoking amount in the model was found to be less than 2.0×10^-16^, indicating its predictive power. Based on these findings, the decision was made to use the smoking amount (packs×years) as the clinical factor for subsequent modeling.

After the smoking amount was included in the LR model, AUROCs of 94% and 93% were achieved using the modified LR model for the discovery and validation cohorts, respectively (Fig 2D). The Youden index of the discovery AUROC curve achieved 85% sensitivity and 88% specificity. AUROCs of 97% and 97% were achieved by the precision-recall curve of the model for the discovery and validation cohorts, respectively (Fig 2G). When the smoking amount was included, the previously reported discovery-AUROC/validation-AUROC values were increased to 94%/92% and 94%/89%, for the early-stages patients (Stage I and II) and the Stage I patients, respectively (Fig 2E and Fig 2F). The sensitivity and specificity of the LR model for the early-stages patients were 88% and 81%, respectively (Youden Index). For the Stage I patients, the sensitivity and specificity of 91% and 80%, respectively, were reached by the LR model (Youden Index). As shown in Fig 2H, the area under the precision-recall curves of the early-stage patients’ LR model were 96% for both the discovery set and the validation set. For the Stage I patients, 95% and 90% of the precision-recall curve AUROC values were achieved by the LR model, for the discovery set and the validation set, respectively (Fig 2I). Other details of the three models described above are listed in Table 2 to 4.

**Table 2.**
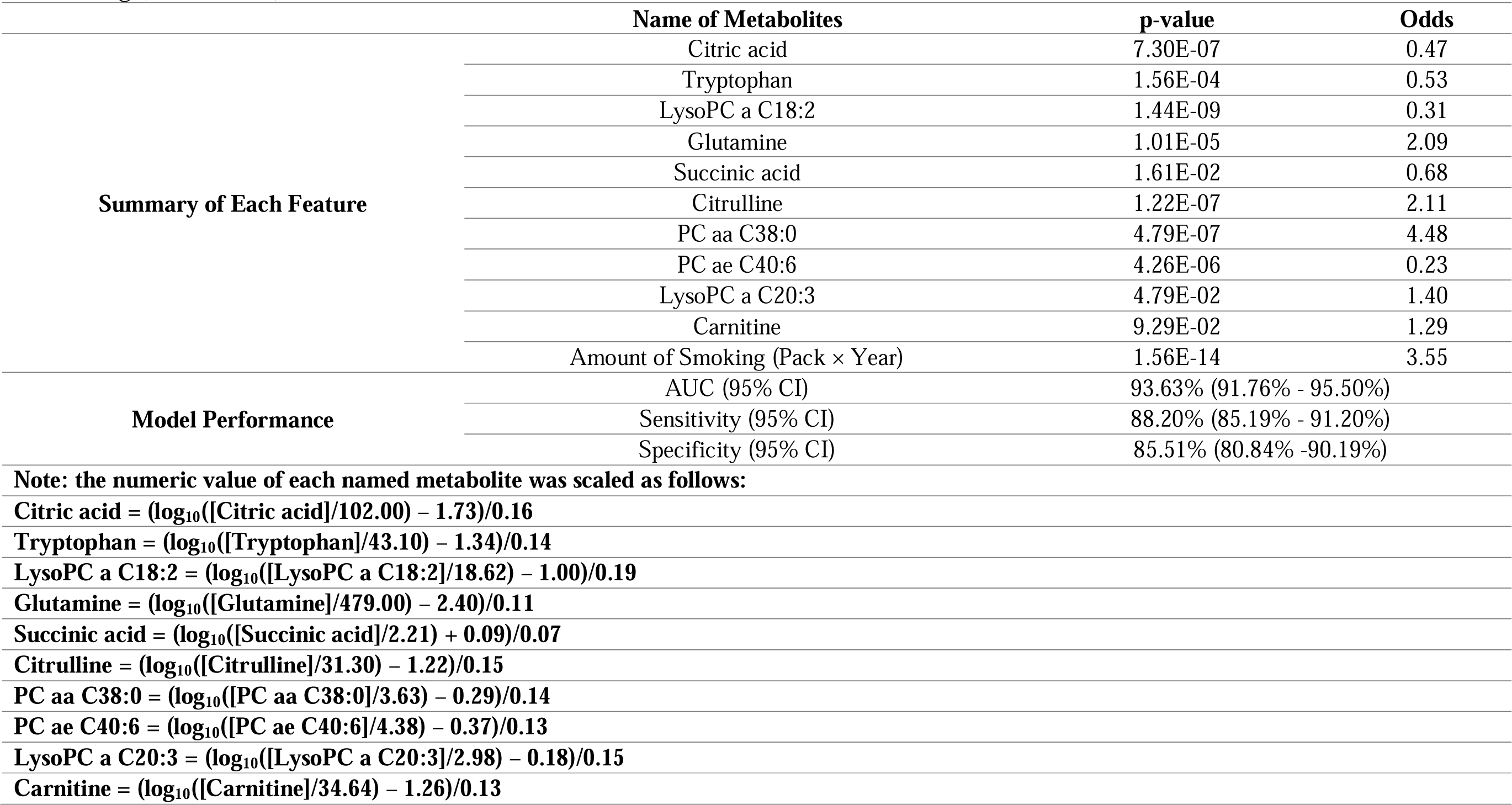
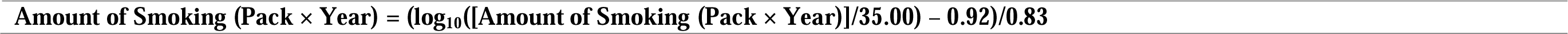
The logistic regression-based optimal model for NSCLC patients vs controls. Values in square brackets represent measured (unscaled) concentrations of the metabolites. Values in square brackets represent measured (unscaled) concentrations of the metabolitesor unscaled amount of smoking (Pack × Year).

**Table 3.**
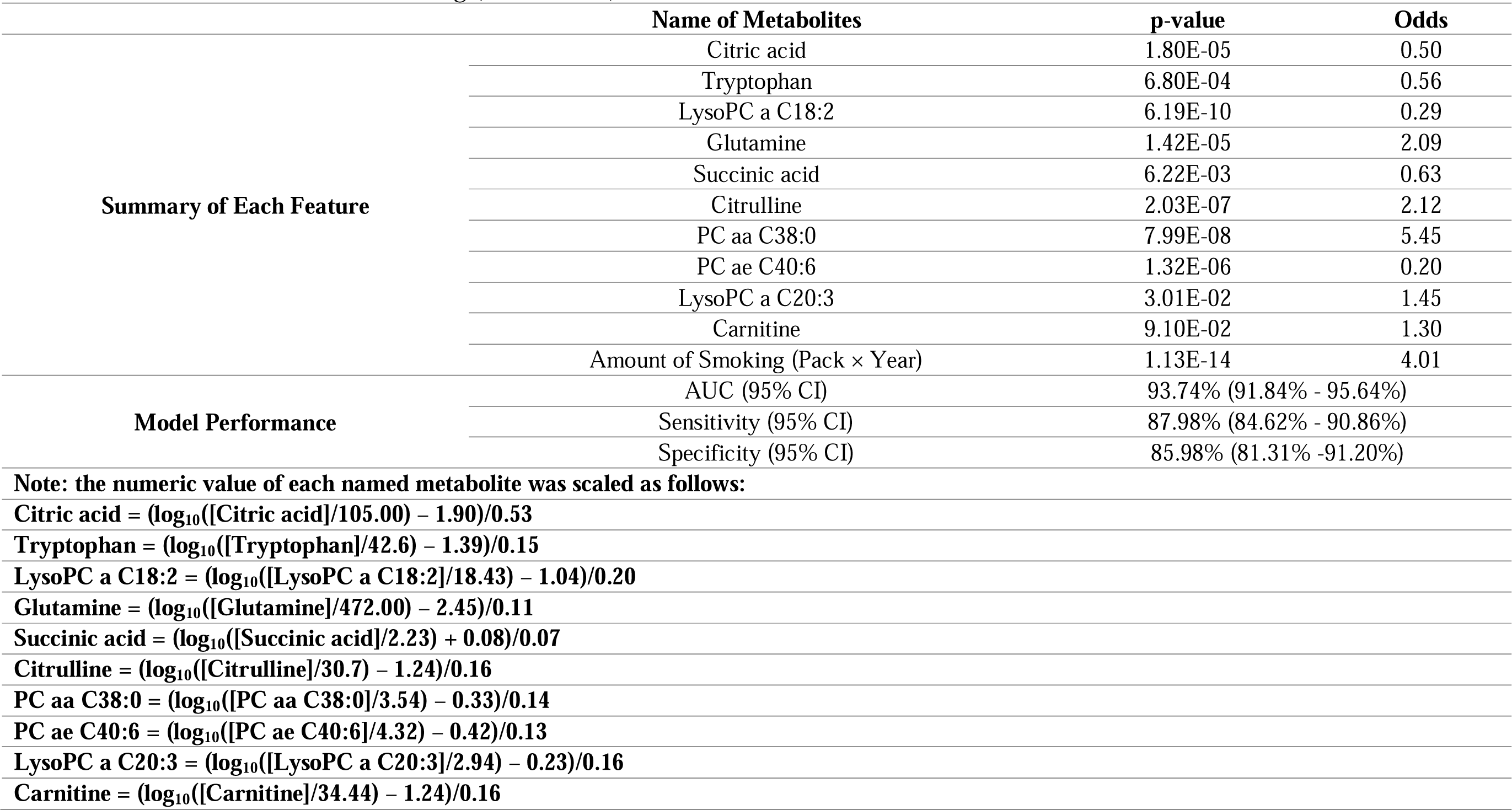
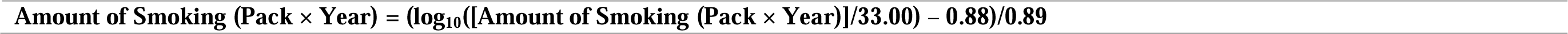
The logistic regression-based optimal model for early-stage (Stage I + Stage II) NSCLC patients vs controls. Values in square brackets represent measured (unscaled) concentrations of the metabolites. Values in square brackets represent measured (unscaled) concentrations of the metabolites or unscaled amount of smoking (Pack × Year).

**Table 4.**
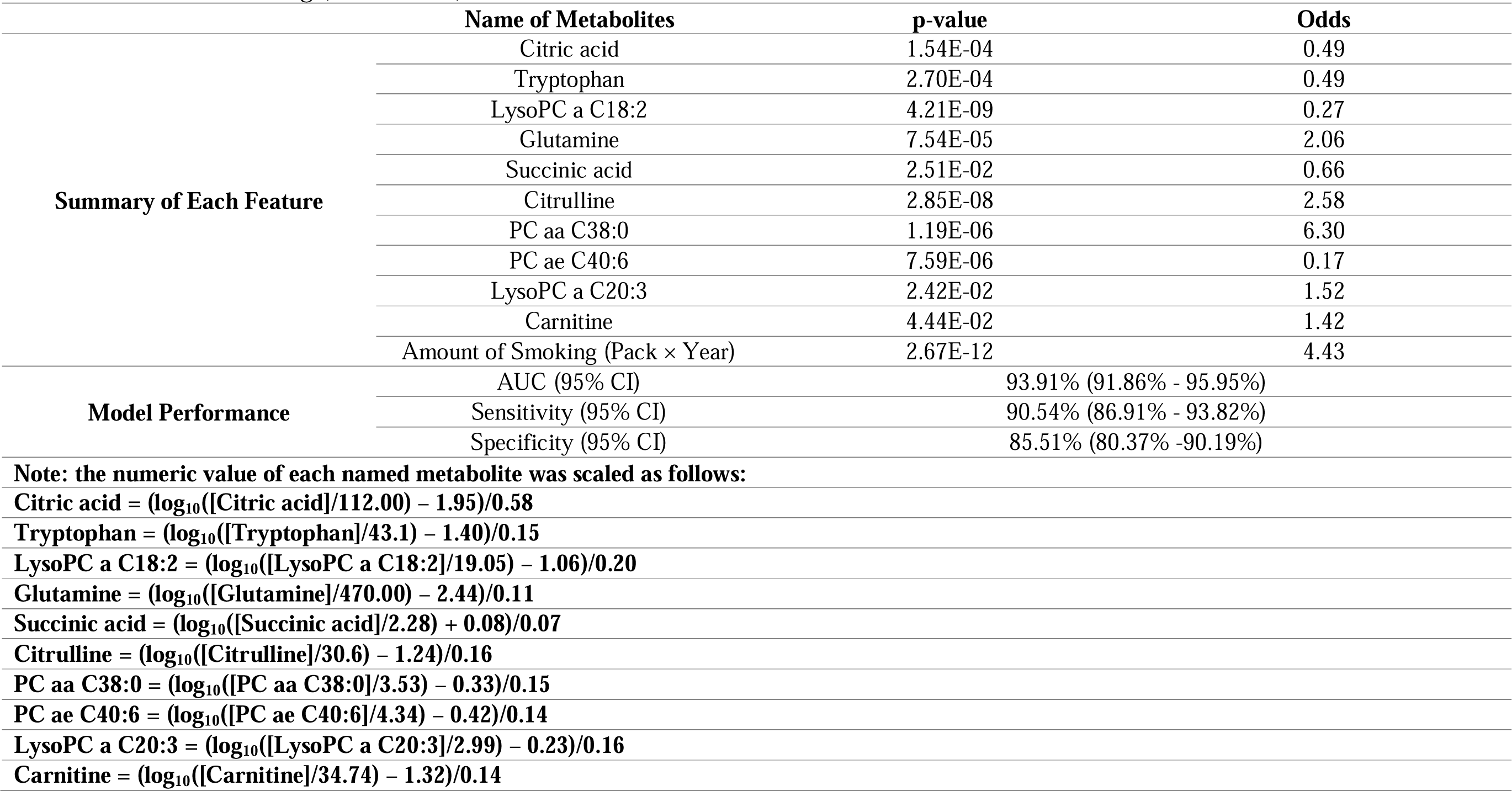
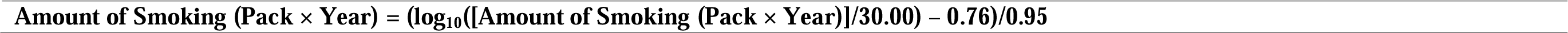
The logistic regression-based optimal model for Stage I NSCLC patients vs controls. Values in square brackets represent measured (unscaled) concentrations of the metabolites. Values in square brackets represent measured (unscaled) concentrations of the metabolites or unscaled amount of smoking (Pack × Year).

## Discussion

In both our previous and current studies, we applied quantitative LC-MS/MS-based metabolomics techniques to gain valuable insights into the metabolic alterations associated with NSCLC. The use of absolute or semi-quantitative data in statistical analyses and modeling allowed us to conduct more precise analyses and achieve more consistent outcomes regarding alterations in the metabolomic profiles of individuals with NSCLC. Previously, we demonstrated that high-performance LR models could distinguish NSCLC patients from healthy individuals using the blood levels of β-hydroxybutyric acid, lysoPC 20:3, PC ae C40:6, citric acid, and fumaric acid. In this study, we applied the same quantitative techniques to a much larger sample from the same population to validate our previous findings. When we built the LR model using only the five previously published metabolites and the smoking frequency (measured as the amount of pack cigarettes smoked per year), the AUROC of the discovery set reached 87% (data not shown). This result indicates that our previous study, despite its smaller sample size, correctly identified metabolomic differences in early-stage NSCLC patients compared to the non-cancer population.

To achieve a model with superior performance that fits both the discovery cohort and validation cohort, we conducted multiple statistical analyses and LR modeling. Ultimately, we selected ten metabolites to build a high-performance predictive model for early-stage NSCLC. β-hydroxybutyric acid and fumaric acid were excluded from the new model, while lysoPC 20:3, PC ae C40:6, and citric acid remained as significant predictive components. Of the ten metabolites selected for the model, two were from both the lysoPC and PC families. Decreased levels of polyunsaturated PCs have been previously reported in lung cancer (22), and altered lysoPC levels have also been documented in various cancer studies (23,24). Although no definitive conclusions have been drawn regarding the relationship between lung cancer and these two lipid families, the consistent changes observed for lysoPC a C20:3, lysoPC a C18:2, PC ae 40:6, and PC aa C38:0 in both the discovery and validation sets strongly suggests that changes in these lipids families may be a characteristic of the blood metabolome in early-stage NSCLC patients.

In addition to these four lipids, the remaining six small-molecule metabolites included in the model have been widely associated with tumors (25–35). Alterations in the concentrations of these metabolites in the blood of NSCLC patients may be related to the metabolic reprogramming of tumor tissues. We also examined the distribution of clinical variables such as age, smoking history, sex, race, height, weight, and BMI, among NSCLC patients and controls. Of the collected samples, only age and smoking history showed significant differences between NSCLC patients and controls. We then further explored the role of age and smoking history on the ability to discriminate NSCLC patients from controls. While the inclusion of age did not significantly affect the model’s performance, the addition of smoking history did improve it. Thus, the final model used metabolites and smoking frequency as biomarkers to differentiate between NSCLC and control groups.

This study minimized the influence of clinical variables on the metabolomics profiles, as the samples were balanced across groups for sex, race, and BMI. Although age and BMI did not significantly enhance the model’s performance, other studies have shown a significant association between these factors and the risk of NSCLC (36–38). Future follow-up studies in different cohorts are necessary to explore the potential of incorporating BMI and age into the modeling to improve diagnostic accuracy. Additionally, lifestyle and environmental exposure factors are closely related to the risk of NSCLC. In subsequent studies, it will be important to consider how to integrate these factors into the prediction model to further improve its performance in real-world situations.

In this study, the control group included patients with non-cancerous lung diseases such as COPD, asthma, benign lung tumors, and bronchitis. These patients may exhibit symptoms similar to early-stage lung cancer, potentially complicating the clinical diagnosis of lung cancer. Despite this, our predictive model remained robust highlighting its potential applicability in real-world lung cancer screening scenarios. Our findings also indicate significant differences in the metabolomic profiles between NSCLC and non-cancerous lung diseases. It is worth noting that patients with non-cancerous lung diseases accounted for 63% of the subjects enrolled in the control group. To ensure the transferability of our findings, future studies should validate our results using samples that reflect the distribution of patients with non-cancerous lung diseases in the target population of lung cancer screening programs.

The metabolomics-based model developed in thus study holds significant clinical potential for early detection of NSCLC, especially when compared to currently available methods such as LDCT and liquid biopsy techniques like ctDNA assays. LDCT, while widely used for lung cancer screening, presents several limitations including low specificity (61-73%), which often results in false positives and unnecessary follow-up procedures (39–42). Our metabolomic model, by comparison, demonstrated much higher AUROC (above 90%) for early-stage NSCLC, suggesting improved specificity and fewer false positives. This makes it a strong candidate for its integration into existing screening programs, possibly reducing the burden of unnecessary diagnostic interventions.

On the other hand, ctDNA assays, while offering a non-invasive alternative, struggle with low sensitivity (50-70%) in detecting early-stage lung cancer. In contrast, our model’s ability to detect metabolic changes associated with early-stage NSCLC makes it a more sensitive and robust method for early detection. By focusing on the metabolic reprograming characteristics of NSCLC, our approach provides an advantage over ctDNA, which may need higher levels of circulating tumor material (found in later disease stages) for accurate detection (43–45) .

Overall, the integration of our metabolomics-based method with traditional techniques could improve the accuracy and efficiency of lung cancer screening programs. The relatively low cost, minimal invasiveness, and high sensitivity of our method make it an appealing addition to clinical workflows, particularly for high-risk populations.

## Conclusion

In summary, this study validates the use of large-scale, targeted quantitative metabolomics analysis for identifying diagnostic metabolic biomarkers of early-stage NSCLC. Our LR model performed remarkably well in distinguishing stage I and stage II NSCLC patients from the control group, with an AUROC exceeding 90% in both the discovery and validation sets. These results underscore each of the models’ predictive strengths and clinical relevance.

These promising results pave the way for the development of a minimally invasive, highly efficient, scalable, and cost-effective lung cancer screening assay. This assay, which would require as little as 10 μL of plasma and which could be processed within minutes using a standard clinical-grade mass spectrometer, represents a practical solution for widespread clinical adoption. Its affordability and ease of use could enhance accessibility to early lung cancer screening, particularly in resource-limited settings.

In a broader context, our study highlights the significant clinical utility of metabolite biomarkers, especially when combined with a patient’s smoking history, for the early detection of NSCLC. This approach offers promising potential to improve patient outcomes through timely intervention, which could lead to increased survival rates and enhanced quality of life. Furthermore, integrating metabolite biomarkers into routine lung cancer screening could offer a more comprehensive diagnostic strategy, reducing false positives and refining treatment decisions.

Future research should focus on validating these findings across diverse populations and exploring the integration of metabolite biomarkers with emerging technologies such as deep learning to enhance diagnostic precision. Additionally, expanding this approach to monitor disease progression could unlock new possibilities in personalized medicine. The incorporation of metabolomics into routine cancer screening not only represents a significant advancement in the field of oncology but also has the potential to reshape current healthcare practices.

## Data Availability

All data produced in the present study are available upon reasonable request to the authors

## Acknowledgement

The clinical samples used in this study were provided by the Institut universitaire de cardiologie et de pneumologie de Québec (IUCPQ), which is supported by the Quebec Respiratory Health Research Network and the IUCPQ Foundation.

