## Supplementary Materials for "Clinical Validation of Metabolite Markers for Early Lung Cancer Detection"

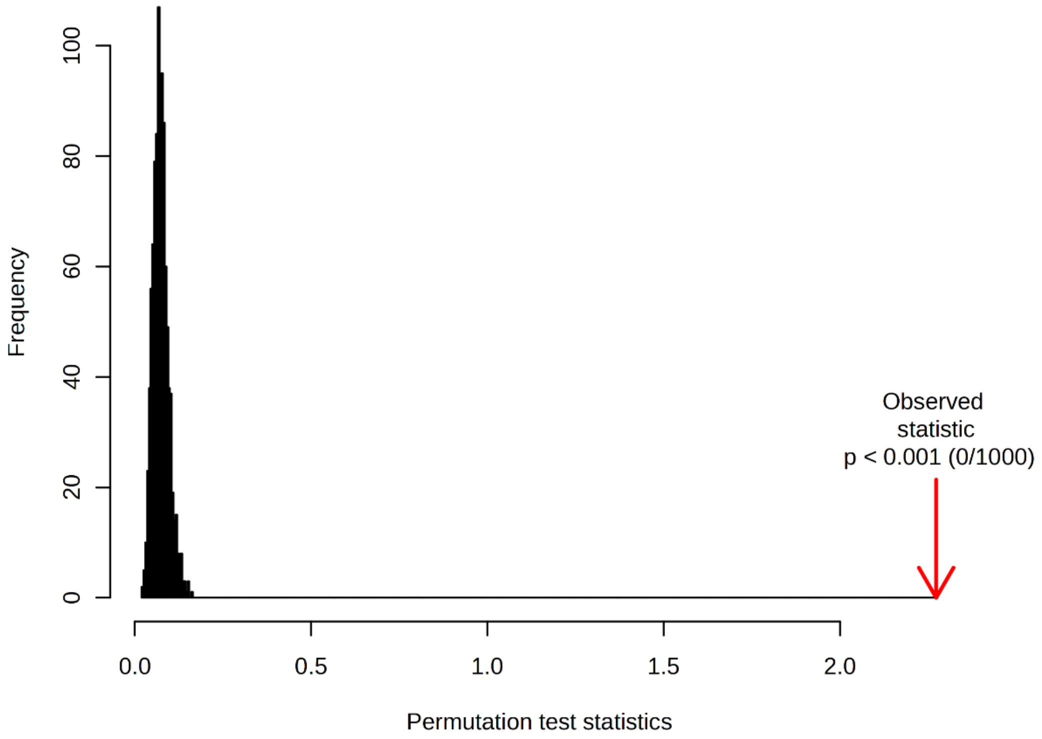


**Figure S1** Permutation tests of the partial least squared- discriminant analyses (PLS-DA): NSCLC at all stages versus controls.

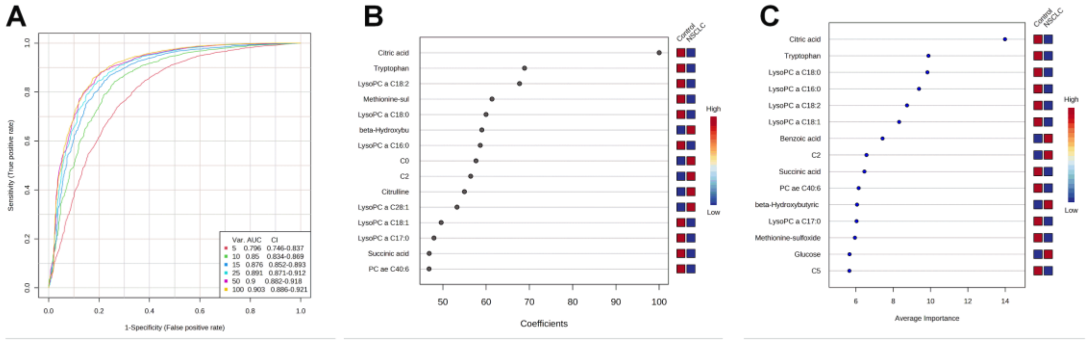


**Figure S2** (A) ROC curves of the random forest exploration models with different numbers of metabolite features for NSCLC at all stages. Number of metabolite features in each model are indicated as Var. in the right-bottom box; (B) Variable importance in projection plot of the PLS-DA for NSCLC at all stages. The most discriminating metabolites are shown in descending order of coefficient scores. The color boxes indicate whether metabolite concentration is increased (red) or decreased (blue) in controls vs. cases. (C) The most frequently selected metabolites (Number of features = 25) in the random forest exploration models. The color boxes indicate whether metabolite concentration is increased (red) or decreased (blue) in controls vs. cases.

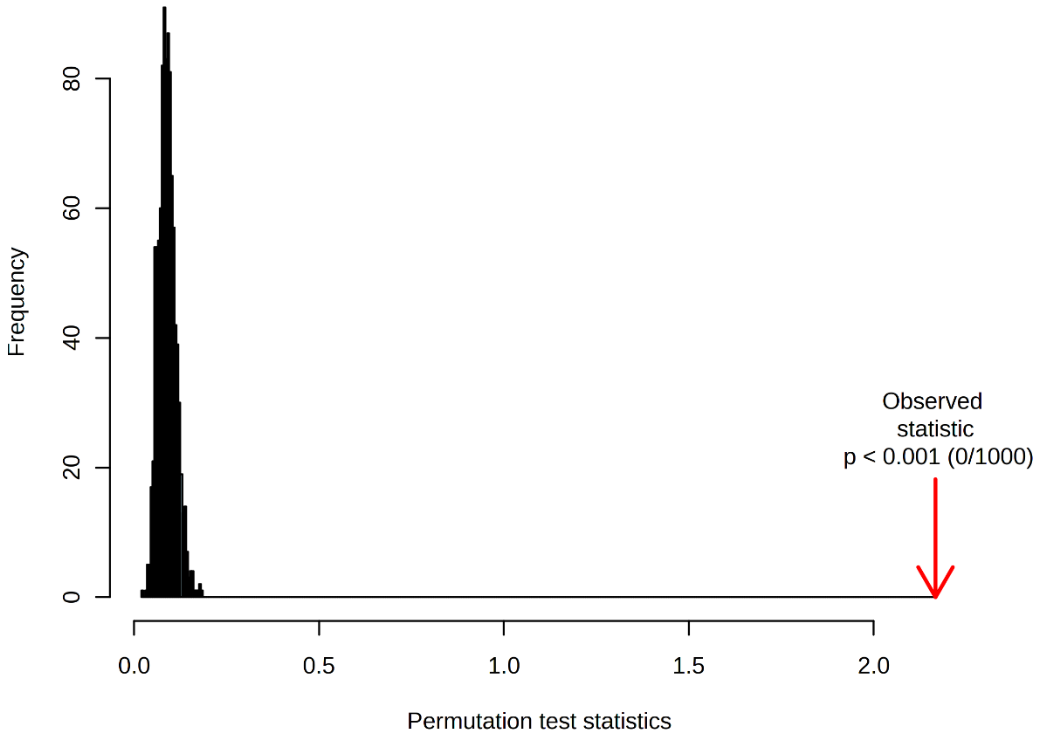


**Figure S3** Permutation tests of the partial least squared- discriminant analyses (PLS-DA): Early-Stage NSCLC (Stage I + Stage II) versus controls.

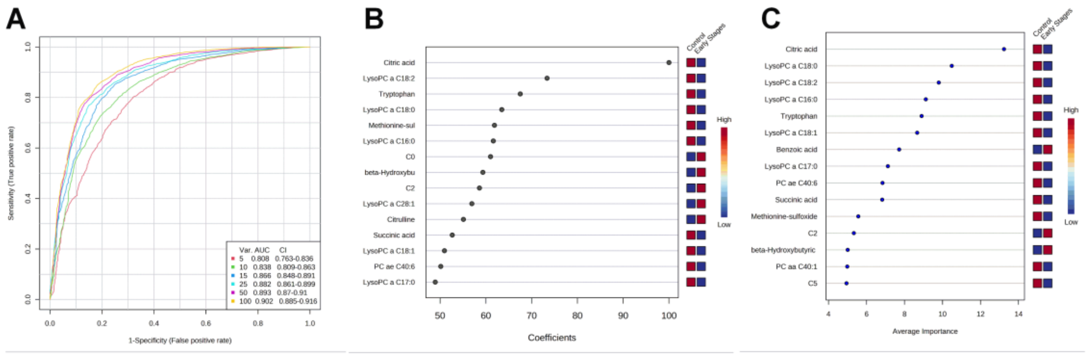


**Figure S4** (A) ROC curves of the random forest exploration models with different numbers of metabolite features for early-stage NSCLC. Number of metabolite features in each model are indicated as Var. in the right-bottom box; (B) Variable importance in projection plot of the PLS-DA for early-stage NSCLC. The most discriminating metabolites are shown in descending order of coefficient scores. The color boxes indicate whether metabolite concentration is increased (red) or decreased (blue) in controls vs. cases. (C) The most frequently selected metabolites (Number of features = 25) in the random forest exploration models. The color boxes indicate whether metabolite concentration is increased (red) or decreased (blue) in controls vs. cases.

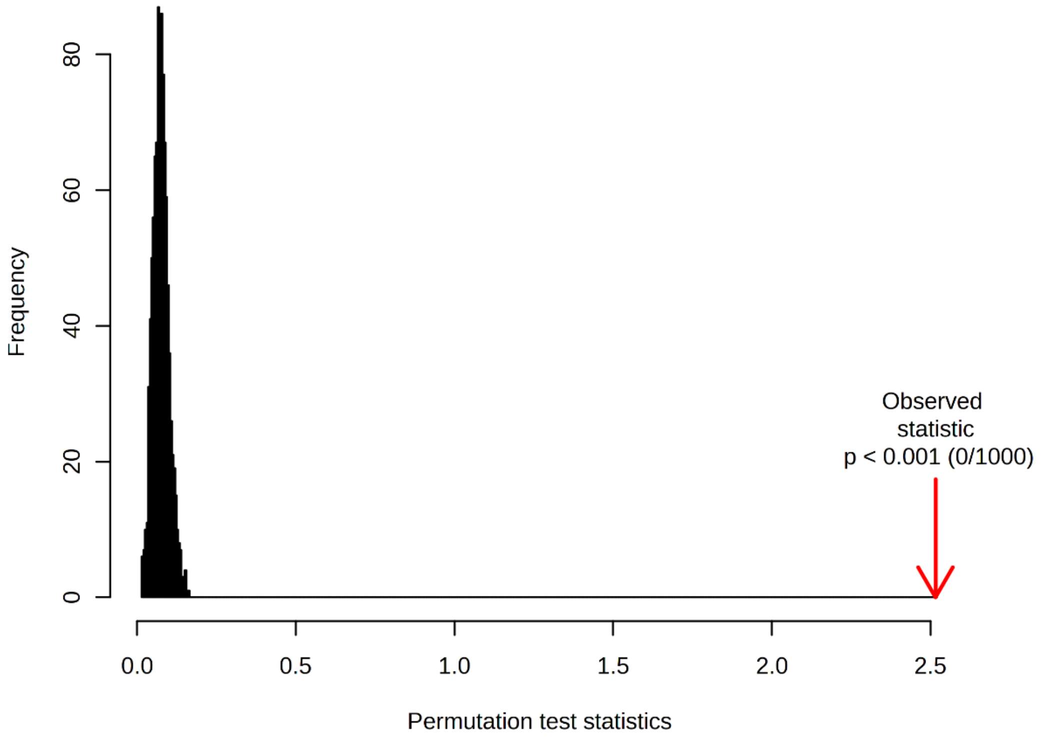


**Figure S5** Permutation tests of the partial least squared- discriminant analyses (PLS-DA): Stage I NSCLC versus controls.

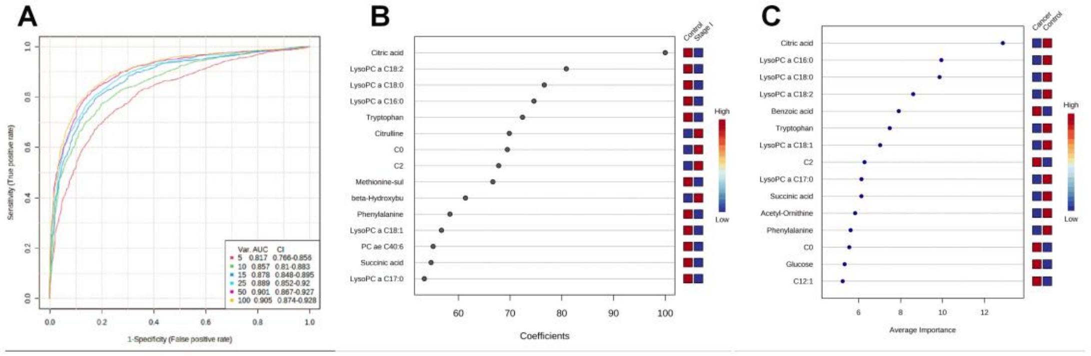


**Figure S6** (A) ROC curves of the random forest exploration models with different numbers of metabolite features for stage I NSCLC. Number of metabolite features in each model are indicated as Var. in the right-bottom box; (B) Variable importance in projection plot of the PLS-DA for stage I NSCLC. The most discriminating metabolites are shown in descending order of coefficient scores. The color boxes indicate whether metabolite concentration is increased (red) or decreased (blue) in controls vs. cases. (C) The most frequently selected metabolites (Number of features = 25) in the random forest exploration models. The color boxes indicate whether metabolite concentration is increased (red) or decreased (blue) in controls vs. cases.

**Table S1.** Summary of the control samples.

| Discovery Set | | | | | | | | | | |
|---|---|---|---|---|---|---|---|---|---|---|
| **Group** | Number of Samples | Age | | | Sex | | Smoking status | | | |
| | | Range | Mean | Median | Male | Female | Never | Former | Current | Median Pack × Years (Former + Current) |
| **Healthy** | 80 | 40-82 | 59.5 | 59 | 39 | 41 | 46 | 28 | 6 | 15 |
| **Asthma** | 31 | 46-78 | 63.9 | 62 | 16 | 15 | 12 | 15 | 4 | 33 |
| **Benign tumor** | 11 | 28-71 | 54.4 | 54 | 3 | 8 | 1 | 5 | 5 | 19.8 |
| **Bronchiectasis** | 8 | 49-73 | 59.9 | 59 | 4 | 4 | 3 | 3 | 2 | 35 |
| **COPD** | 46 | 47-89 | 61.3 | 58 | 24 | 22 | 8 | 28 | 10 | 33.1 |
| **COVID** | 38 | 39-90 | 63.3 | 62 | 25 | 13 | 28 | 9 | 1 | 35 |
| **Total** | 214 | 28-90 | 63.5 | 64 | 111 | 103 | 98 | 88 | 28 | 34 |

| Validation Set | | | | | | | | | | |
|---|---|---|---|---|---|---|---|---|---|---|
| **Group** | Number of Samples | Age | | | Sex | | Smoking status | | | |
| | | Range | Mean | Median | Male | Female | Never | Former | Current | Median Pack × Years (Former + Current) |
| **Healthy** | 60 | 49-79 | 66.1 | 67 | 26 | 44 | 14 | 40 | 16 | 30 |

**Table S2**. Metabolites with significant different between the NSCLC patients (at all stages) and the controls using univariate statistical analysis (Mann Whitney Rank Sum test).

| Name of Metabolites | Fold Change (Case/Control) | p-value |
|---|---|---|
| **Citric acid** | 0.749 | 8.27E-27 |
| Benzoic acid | 1.908 | 1.24E-18 |
| LysoPC a C18:2 | 0.767 | 1.70E-16 |
| beta-Hydroxybutyric acid | 2.056 | 2.27E-16 |
| LysoPC a C18:0 | 0.803 | 2.48E-16 |
| C2 | 1.235 | 2.68E-16 |
| Tryptophan | 0.828 | 3.77E-15 |
| LysoPC a C16:0 | 0.822 | 3.30E-14 |
| Citrulline | 1.192 | 9.11E-14 |
| Methionine-sulfoxide | 0.757 | 2.50E-12 |
| LysoPC a C18:1 | 0.832 | 1.19E-11 |
| C0 | 1.116 | 1.34E-08 |
| LysoPC a C17:0 | 0.859 | 2.48E-08 |
| Glycine | 1.144 | 9.41E-08 |
| Succinic acid | 0.771 | 1.54E-07 |
| Glutamic acid | 1.166 | 9.21E-07 |
| C5 | 0.858 | 2.12E-06 |
| Asymmetric?dimethylarginine | 1.100 | 2.64E-06 |
| PC ae C40:6 | 0.898 | 2.92E-06 |
| Putrescine | 1.153 | 5.59E-06 |
| Acetyl-Ornithine | 0.747 | 6.65E-06 |
| LysoPC a C28:1 | 1.120 | 1.28E-05 |
| p-Hydroxyhippuric acid | 1.341 | 2.16E-05 |
| Hippuric acid | 1.251 | 2.47E-05 |
| Pyruvic acid | 0.868 | 5.12E-05 |
| LysoPC a C20:3 | 1.109 | 1.12E-04 |
| LysoPC a C26:0 | 1.119 | 2.94E-04 |
| Phenylalanine | 0.880 | 3.58E-04 |
| trans-Hydroxyproline | 1.108 | 4.56E-04 |
| Arginine | 1.132 | 4.65E-04 |
| LysoPC a C26:1 | 1.124 | 5.17E-04 |
| Lactic acid | 0.903 | 5.60E-04 |
| alpha-Aminoadipic acid | 0.525 | 5.73E-04 |
| Aspartic acid | 1.122 | 1.07E-03 |
| HPHPA | 1.386 | 1.98E-03 |
| C7DC | 1.234 | 4.29E-03 |
| Diacetylspermine | 1.212 | 5.99E-03 |
| Cadaverine | 1.403 | 6.46E-03 |
| Glucose | 0.520 | 9.53E-03 |
| Trimethylamine N-oxide | 0.785 | 1.08E-02 |
| Homovanillic acid | 0.379 | 3.21E-02 |

**Table S3**. Metabolites with significant different between the early-stages NSCLC patients and the controls using univariate statistical analysis (Mann Whitney Rank Sum test).

| Name of Metabolites | Fold Change (Case/Control) | p-value |
|---|---|---|
| **Citric acid** | 0.753 | 1.02E-24 |
| LysoPC a C18:2 | 0.757 | 7.82E-17 |
| Benzoic acid | 1.826 | 8.47E-16 |
| LysoPC a C18:0 | 0.807 | 2.14E-15 |
| C2 | 1.216 | 9.49E-15 |
| beta-Hydroxybutyric acid | 1.892 | 3.03E-14 |
| Tryptophan | 0.839 | 6.18E-13 |
| LysoPC a C16:0 | 0.828 | 8.67E-13 |
| Citrulline | 1.186 | 1.29E-12 |
| Methionine-sulfoxide | 0.764 | 3.08E-11 |
| LysoPC a C18:1 | 0.836 | 6.96E-11 |
| C0 | 1.109 | 2.76E-08 |
| Succinic acid | 0.757 | 1.33E-07 |
| LysoPC a C17:0 | 0.867 | 3.10E-07 |
| Glycine | 1.135 | 5.89E-07 |
| LysoPC a C28:1 | 1.132 | 2.87E-06 |
| PC ae C40:6 | 0.896 | 4.47E-06 |
| Glutamic acid | 1.151 | 6.09E-06 |
| C5 | 0.866 | 1.37E-05 |
| Acetyl-Ornithine | 0.756 | 1.50E-05 |
| Pyruvic acid | 0.865 | 3.35E-05 |
| Putrescine | 1.141 | 4.01E-05 |
| Hippuric acid | 1.243 | 1.28E-04 |
| LysoPC a C20:3 | 1.109 | 1.43E-04 |
| p-Hydroxyhippuric acid | 1.308 | 1.55E-04 |
| LysoPC a C26:0 | 1.123 | 2.31E-04 |
| Phenylalanine | 0.876 | 2.53E-04 |
| Lactic acid | 0.899 | 3.74E-04 |
| LysoPC a C26:1 | 1.128 | 4.58E-04 |
| trans-Hydroxyproline | 1.110 | 7.67E-04 |
| HPHPA | 1.395 | 1.76E-03 |
| Aspartic acid | 1.117 | 2.04E-03 |
| Arginine | 1.105 | 2.19E-03 |
| C7DC | 1.261 | 4.88E-03 |
| alpha-Aminoadipic acid | 0.563 | 6.99E-03 |
| Trimethylamine N-oxide | 0.808 | 1.64E-02 |
| Diacetylspermine | 1.143 | 1.70E-02 |
| Cadaverine | 1.184 | 1.88E-02 |
| Homovanillic acid | 0.374 | 2.17E-02 |
| Glucose | 0.516 | 3.42E-02 |

**Table S4.** Metabolites with significant different between the stage I NSCLC patients and the controls using univariate statistical analysis (Mann Whitney Rank Sum test).

| Name of Metabolites | Fold Change (Case/Control) | p-value |
|---|---|---|
| **Citric acid** | 0.663 | 8.78E-19 |
| LysoPC a C18:2 | 0.751 | 9.86E-15 |
| Benzoic acid | 1.772 | 2.24E-14 |
| LysoPC a C18:0 | 0.798 | 2.30E-14 |
| LysoPC a C16:0 | 0.818 | 9.93E-13 |
| C2 | 1.209 | 1.87E-12 |
| Citrulline | 1.199 | 1.20E-11 |
| beta-Hydroxybutyric acid | 1.684 | 7.13E-10 |
| Tryptophan | 0.852 | 1.32E-09 |
| Methionine-sulfoxide | 0.775 | 9.19E-09 |
| LysoPC a C18:1 | 0.843 | 1.26E-08 |
| C0 | 1.110 | 8.10E-08 |
| LysoPC a C17:0 | 0.867 | 2.03E-06 |
| Pyruvic acid | 0.830 | 4.77E-06 |
| PC ae C40:6 | 0.893 | 1.03E-05 |
| Acetyl-Ornithine | 0.755 | 1.16E-05 |
| Succinic acid | 0.761 | 1.63E-05 |
| Glycine | 1.111 | 2.07E-05 |
| Putrescine | 1.136 | 5.56E-05 |
| Phenylalanine | 0.859 | 8.80E-05 |
| Glutamic acid | 1.141 | 1.85E-04 |
| Lactic acid | 0.882 | 4.18E-04 |
| C5 | 0.874 | 5.61E-04 |
| trans-Hydroxyproline | 1.124 | 5.75E-04 |
| C7DC | 1.294 | 2.06E-03 |
| p-Hydroxyhippuric acid | 1.256 | 2.54E-03 |
| Hippuric acid | 1.217 | 3.01E-03 |
| Isobutyric acid | 1.103 | 3.06E-03 |
| HPHPA | 1.423 | 4.02E-03 |
| Aspartic acid | 1.115 | 4.73E-03 |
| Homovanillic acid | 0.364 | 6.62E-03 |
| Methionine | 0.899 | 7.63E-03 |
| alpha-Aminoadipic acid | 0.586 | 4.00E-02 |
